# Seeding COVID-19 across sub-Saharan Africa: an analysis of reported importation events across 48 countries

**DOI:** 10.1101/2020.04.01.20050203

**Authors:** Laura A Skrip, Prashanth Selvaraj, Brittany Hagedorn, Andre Lin Ouédraogo, Navideh Noori, Dina Mistry, Jamie Bedson, Laurent Hébert-Dufresne, Samuel V Scarpino, Benjamin M Althouse

## Abstract

**Background:** The first case of COVID-19 in sub-Saharan Africa (SSA) was reported by Nigeria on February 27, 2020. While case counts in the entire region remain considerably less than those being reported by individual countries in Europe, Asia, and the Americas, SSA countries remain vulnerable to COVID morbidity and mortality due to systemic healthcare weaknesses, less financial resources and infrastructure to address the new crisis, and untreated comorbidities. Variation in preparedness and response capacity as well as in data availability has raised concerns about undetected transmission events.

**Methods:** Confirmed cases reported by SSA countries were line-listed to capture epidemiological details related to early transmission events into and within countries. Data were retrieved from publicly available sources, including institutional websites, situation reports, press releases, and social media accounts, with supplementary details obtained from news articles. A data availability score was calculated for each imported case in terms of how many indicators (sex, age, travel history, date of arrival in country, reporting date of confirmation, and how detected) could be identified. We assessed the relationship between time to first importation and overall Global Health Security Index (GHSI) using Cox regression. K-means clustering grouped countries according to healthcare capacity and health and demographic risk factors.

**Results:** A total of 13,201 confirmed cases of COVID-19 were reported by 48 countries in SSA during the 54 days following the first known introduction to the region. Out of the 2516 cases for which travel history information was publicly available, 1129 (44.9%) were considered importation events. At the regional level, imported cases tended to be male (65.0%), were a median 41.0 years old (Range: 6 weeks - 88 years), and most frequently had recent travel history from Europe (53.1%). The median time to reporting an introduction was 19 days; a country’s time to report its first importation was not related to GHSI, after controlling for air traffic. Countries that had, on average, the highest case fatality rates, lowest healthcare capacity, and highest probability of premature death due to non-communicable diseases were among the last to report any cases.

**Conclusions:** Countries with systemic, demographic, and pre-existing health vulnerabilities to severe COVID-related morbidity and mortality are less likely to report any cases or may be reporting with limited public availability of information. Reporting on COVID detection and response efforts, as well as on trends in non-COVID illness and care-seeking behavior, is critical to assessing direct and indirect consequences and capacity needs in resource-constrained settings. Such assessments aid in the ability to make data-driven decisions about interventions, country priorities, and risk assessment.

**Key Messages:**

- We line-listed epidemiological indicators for the initial cases reported by 48 countries in sub-Saharan Africa by reviewing and synthesizing information provided by official institutional outlets and news sources.
- Our findings suggest that countries with the largest proportions of untreated comorbidities, as measured by probability of premature death due to non-communicable diseases, and the fewest healthcare resources tended to not be reporting any cases at one-month post-introduction into the region.
- Using data availability as a measure of gaps in detection and reporting and relating them to COVID-specific parameters for morbidity and mortality provides a measure of vulnerability.
- Accurate and available information on initial cases in seeding local outbreaks is key to projecting case counts and assessing the potential impact of intervention approaches, such that support for local data teams will be important as countries make decisions about control strategies.

## Introduction

The Severe Acute Respiratory Syndrome coronavirus 2 (SARS-CoV 2) virus has spread feverishly across the globe, causing hundreds of thousands of coronavirus 2019 (COVID-19) cases and tens of thousands of deaths as of late March 2020.^1^ While detection of cases has centered on Asia, Europe, and the United States, there so far seems to be a paucity of cases across the continent of Africa, despite regular air traffic in and out, especially that resulting from strong economic and development ties with China.^2^ This lack of cases may be due to inadequate testing capacity.^3^ It is critical to understand how SARS-CoV2 is introduced into countries^45^ in order to anticipate onward transmission of the virus and subsequent risk of infection among population subgroups most vulnerable to severe morbidity and mortality.^6^

Africa may be uniquely positioned to have the most severe and under-detected outcomes related to COVID-19 infection.^7^ The continent’s countries are among those most at-risk of widespread disease threats, per several indices of epidemic preparedness. The World Health Organization’s (WHO) State Party Self-Assessment Annual Reporting (SPAR) database assigns scores to countries to assess capacities needed to detect, assess, notify, report, and respond to public health risk and acute events of domestic and international concern.^5,8^ Similarly, the Infectious Disease Vulnerability Index (IDVI) developed by the RAND Corporation and the Global Health Security Index (GHSI) by Johns Hopkins University ^9^ use a variety of healthcare, economic, demographic, and political factors to assess the vulnerability of a country to prevent or contain an infectious disease outbreak.^10^ Using such indices, recent work has shown most of Sub-Saharan Africa (SSA) to be at risk of COVID-19 importation and at reduced capacity to contain outbreaks due to lack of economic and medical resources.^5^ While it appears that the age groups at highest risk of severe COVID-19 disease and death (those >60 years old)^6,11^ may be proportionately less in many SSA countries than in other parts of the world, the populations in many of these countries are at increased risk of having untreated chronic conditions due to weak health systems.^12^ As a result, individuals with cardiovascular diseases or diabetes,^13,14^ sickle-cell disease, or conditions associated with immunosuppression,^15,16^ which exacerbate the immune response to SARS-CoV2 infection, may contribute to higher-than-expected mortality for younger age groups.

Within the first eight weeks of the first introduction into the region on February 27, 2020, imported seeding events have occurred almost universally in SSA; however, capacity for detection, reporting, and control efforts varies.^5,9^ Across the region and with ranging degrees of enforcement,^17,18^ countries have implemented suites of preventative interventions, including school closures,^19^ curfews, and other social distancing measures,^20^ as well as border and airport closures.^21^ Countries reporting high numbers of cases since the original seeding in SSA were hypothesized to have stronger detection/preparedness systems, such as South Africa and Rwanda. Lower observed case counts or delayed reporting of initial cases, relative to the date of first seeding in SSA, could be due to poor detection. It is the goal of this work to systematically collect and present information on COVID-19 cases in all affected countries in SSA during the first several weeks since known importation, with the aim of giving starting points for prediction of onward transmission. We present reported information in the context of countries’ pandemic preparedness and COVID-specific vulnerability to underscore the risk of undetected transmission -- and therefore significant morbidity and mortality -- in countries with low numbers of recorded cases to date.

## Methods

A line list of confirmed cases in 50 SSA countries (Library of Congress list,^22^ excluding territories) was developed using data published by national governmental institutions *(i.e*., ministries of health and public health institutes) through their websites, press releases, and social media accounts, and supplemented with information from news outlets (Links to all sources are provided in the Supplementary Table). Data on sex, age, travel history (including travel locations and dates of entry into the country where case confirmation occurred), date of reported confirmation, whether a case was due to importation or secondary transmission (both known and community transmission), and information on how the case was detected *(e.g*., active surveillance monitoring or self-presentation) were recorded for each case. We searched using keywords such as ‘COVID’, ‘Ministry of Health’, ‘situation report’, ‘press release’, and/or the date in the language of the respective countries (e.g., English, French, Portuguese, Swahili), although more specific searches in news outlets were done in English. As information was collected from multiple sources, the daily case totals per country in the line list were compared with country-reported totals and/or the WHO situation report totals.^23^ All data collected for the line list is provided as Supplementary Material.

Reported confirmed cases were assumed to be imported when classified as such by national institutions, or when case information included evidence of recent travel history. For countries with limited data available, information on whether cases were imported or due to local transmission was evaluated from the aggregate information in daily WHO situation reports, when feasible. Cases with uncertain travel history or not enough information to determine their importation status were included in the line list but not given a status.

Imported cases were described in terms of sex, 10-year age categories, and time between arrival in country and date that case confirmation was reported. The difference in the sex ratio for imported cases versus for cases due to local transmission was evaluated using a chi-square test (p < 0.05 considered statistically significant). Temporal trends in the frequency of importation events across SSA and in the continents from which they originated were evaluated.

### Preparedness and reporting

The availability of publicly reported data was assessed in terms of the average number of indicators which were reported or could be inferred for each imported case. The indicators included were sex, age, date of case confirmation, travel history, date of arrival in the country, and whether detection was due to active monitoring or self-presentation. Availability was assessed for the first 10 imported cases in each country (or for all cases in countries with fewer than 10 cases reported as of April 21, 2020). To consider changes in availability of individual-level information with increasing incidence, availability was assessed for the second, third, and fourth sets of 10 imported cases in countries with more than 10 reported importation events.

The relationship between country-level pandemic preparedness and case reporting rates was assessed. A Poisson generalized linear model was applied to relate overall GHSI with case counts after adjusting for days since first reported importation as an offset. We also used a Cox proportional hazards model to consider the daily probability of a country reporting any case since the first introduction into the region, after adjusting for the country’s overall GHSI and flight traffic. Flight traffic was included as the annual number of passengers carried by air transport (in billions).^24^ Data for the most recent year available was included for countries with information from 2016 until present. Thirteen out of the 49 SSA countries did not have recent flight traffic data and were excluded from the adjusted Cox model.

### Relationship between risk of severe morbidity and death and reporting

K-means clustering was conducted to identify groups of countries based on risk factors for high rates of severe morbidity and mortality due to COVID-19 or lack of capacity to provide care. Factors included number of healthcare workers per 10,000 population,^25,26^ number of hospital beds per 1,000 population,^27^ proportion of population 60 years or older, and probability of dying between age 30 and exact age 70 from any of cardiovascular disease, cancer, diabetes, or chronic respiratory disease.^28^ The age threshold of 60 years was selected due to evidence to date on increased risk of severe disease in individuals over 60.^6^ Due to limited data available for Western Sahara, it was not included in the cluster analysis. The probability of a country having any case, average data availability score for first 10 importation events, average cumulative reported cases per 100,000 population, and average case fatality as of April 21 were calculated for each of the clusters. Differences in average data availability across and between clusters were evaluated using analysis of variance and the Tukey HSD test.

## Results

### Overview

Out of the 13201 cases listed between February 27 and April 21, 2020, individual-level information on the status *(i.e*., importation or not) for 2516 reported cases could be determined from available information. Among the individuals with known status, 1129 importation events were identified (1129/2516, 44.9%), while 1387 cases were listed as being contacts of known travelers or as a result of unexplained community transmission (1387/2516, 55.1%). Imported cases were majority male (343/528, 65.0%) and were a median 41.0 years old (Range: 6 weeks - 88 years) (Figure 1 & Table 1). Cases due to local transmission were also majority male (177/307, 57.7%) and were a median 35.0 years old (28 days - 105 years). The proportion of males among imported cases was significantly higher than that among cases due to local transmission (p = 0.043). For imported cases, time between arrival into the country and reporting of the confirmed case status ranged from 0 to 40 days (median: 7, IQR: 4-12) (Figure 2).

**Figure 1.**
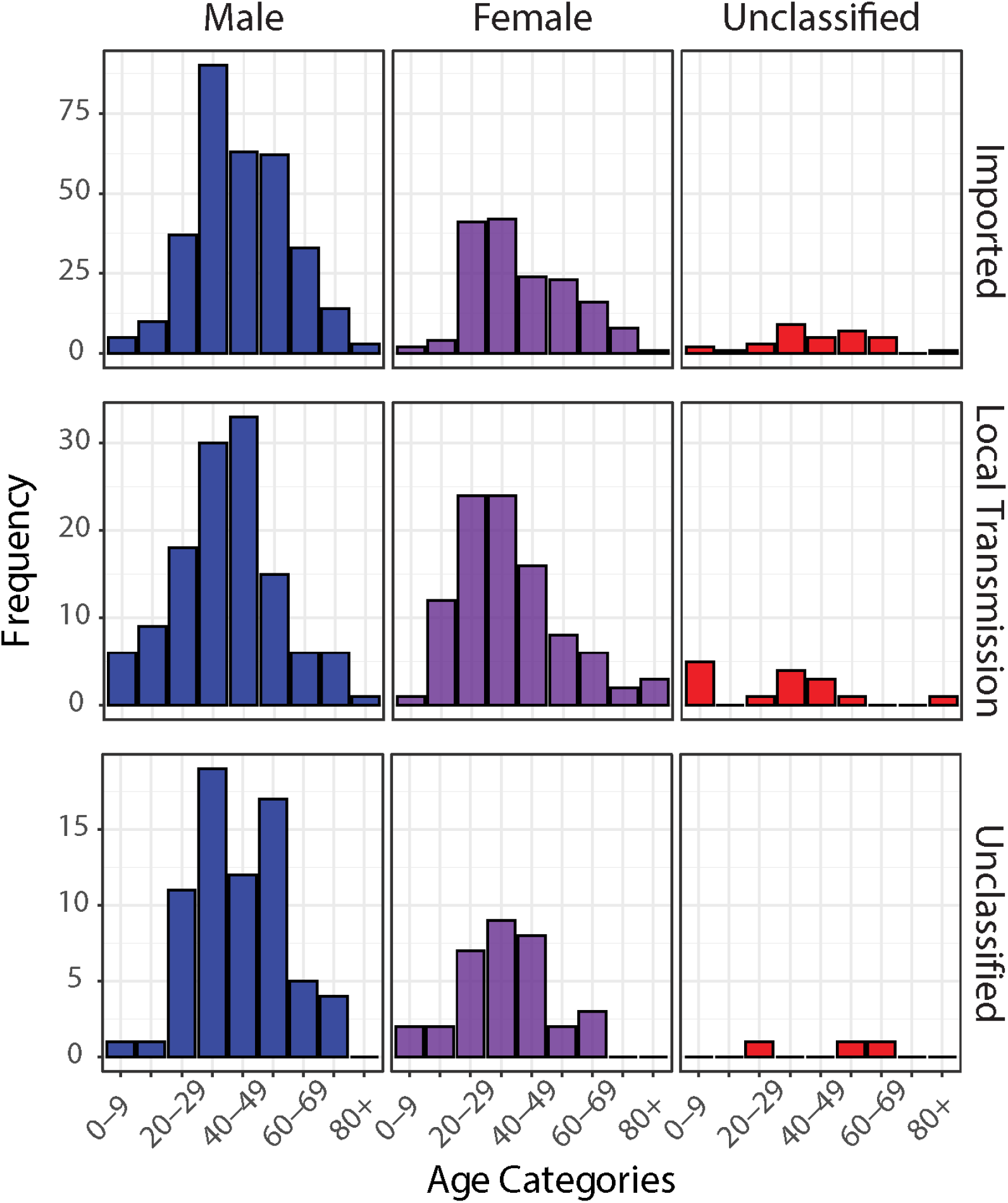
Age-sex distribution of confirmed cases of COVID-19 in sub-Saharan Africa by status (imported cases, cases due to local transmission, and cases with status unavailable). Only cases with both age and sex data are reflected.

**Figure 2.**
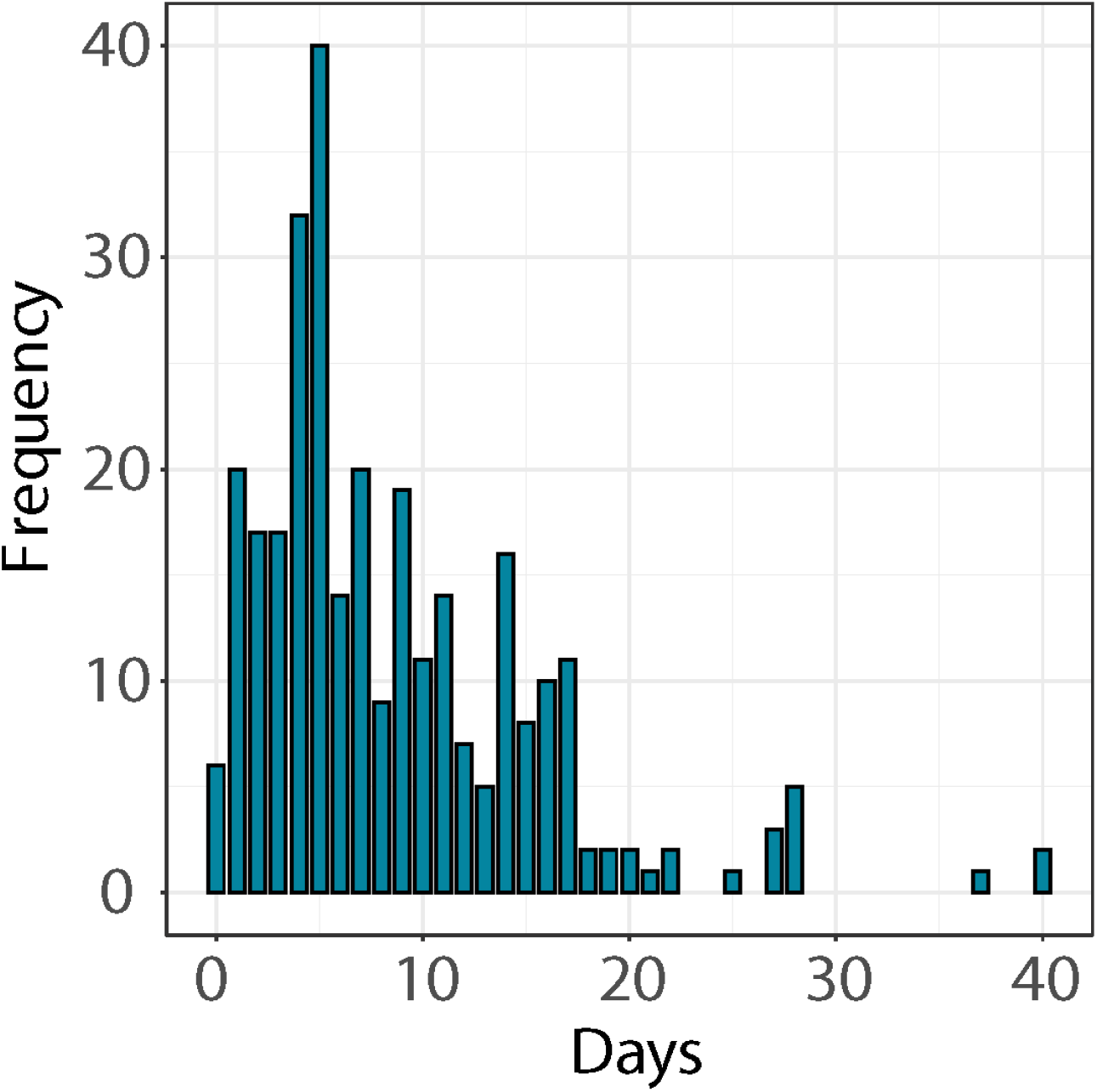
Distribution of time between arrival in country and reporting of case confirmation status

**Table 1.** Characteristics of confirmed case, by status (importation versus local transmission)

| **Characteristic** | **Overall**<br>n=13201 | **Imported**<br>n=1129 | **Local Transmission**<br>n=1387 | **Unknown**<br>n=10685 |
| --- | --- | --- | --- | --- |
| Male | 636 (62.6) | 343 (65.0) | 177 (57.7) | 116 (64.1) |
| Median Age (Range) | 39 years (28 days – 105 years) | 41 years (6 weeks - 88 years) | 35 years (28 days - 105 years) | 39.5 years (2 - 78 years) |
| Mode of Detection |  |  |  |  |
| Active Monitoring | 1327 (93.6) | 446 (89.7) | 483 (93.1) | 396 (99.0) |
| Self-Presentation | 91 (6.4) | 51 (10.3) | 36 (6.9) | 4 (1.0) |

\* Reported as frequency (%) unless otherwise indicated
\*\* Frequencies may not sum to totals per category due to missing information

### Space-time trends

Two SSA countries reported introduction by March 3^rd^ (2/49, 4.1%), eight countries by March 11^th^ (8/49, 16.3%), 31 total countries by March 19^th^ (31/49, 63.3%), and 40 total countries as of March 27^th^ (40/49, 81.6%), and 48 total countries as of April 21^st^ (Figure 3). Comoros and Lesotho remained the only SSA countries not reporting any cases. Among those that could be line-listed, over 50 importation events were each reported by South Africa (198/1129, 17.5%), Senegal (85/1129, 7.5%), Kenya (81/1129, 7.2%), Rwanda (76/1129, 6.7%), Cameroon (71/1129, 6.3%), Ghana (69/1129, 6.1%), and Uganda (53/1129, 4.7%). 60.6% of importation events (684/1129) had travel history available. Most imported cases reported recent travel from Europe (363/684, 53.1%), with fewer reports of travel to Asia (173/684, 25.3%), other affected countries in Africa (92/684, 13.5%), the Americas (52/684, 7.6%), or Oceania (2/684, 0.3%) (Figure 4). Two cases with travel history reported sea travel via a cruise on which they worked (2/684, 0.3%).

**Figure 3.**
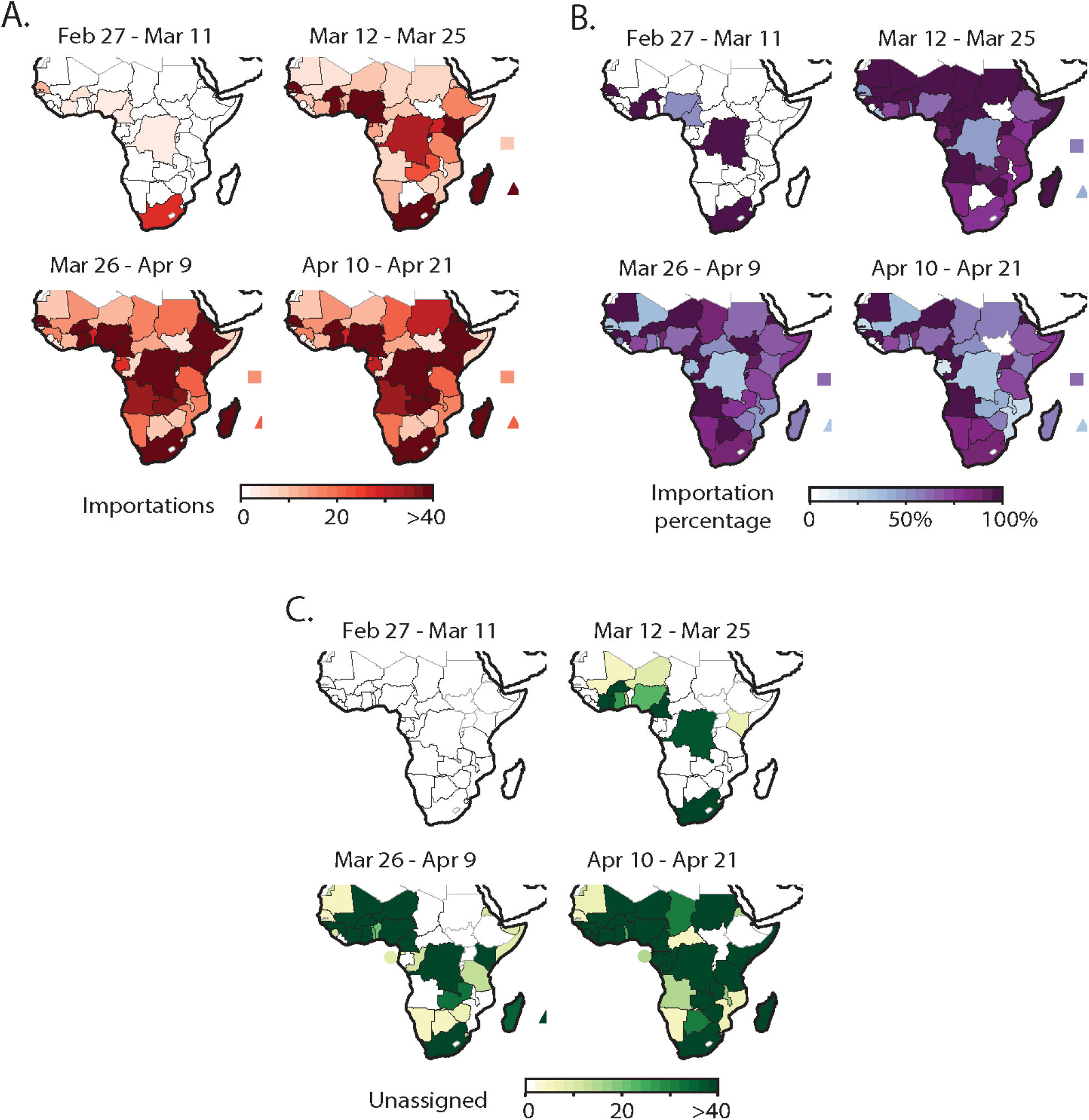
(A) Cumulative numbers of imported cases for which status (importation v. local) is known, by week; (B) Overall percentage of cases (with known status) that are importations; and Panel (C) Number of cases for which status could not be determined from available data. Mauritius and Seychelles are marked by a triangle and square, respectively, given the scale of the maps. Note: In the fourth time period, importation ratios in (B) calculated from the line list were adjusted to reflect aggregate information in three countries for which it was available (South Africa, Senegal, and Mauritius).

**Figure 4.**
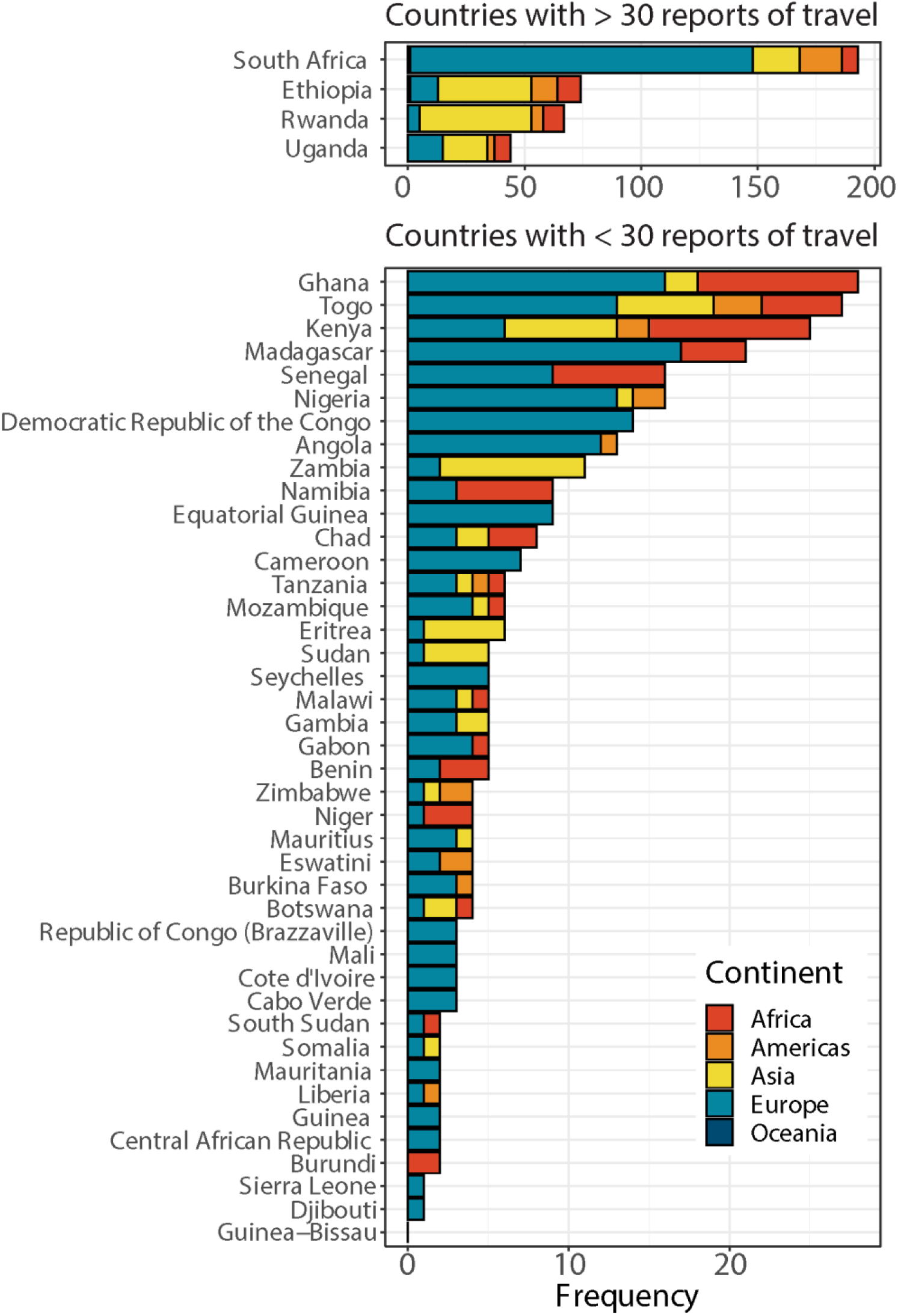
Travel history of imported cases by country of residence and continent of travel, for cases with documented travel history. Multiple entries are shown for confirmed cases with a reported travel itinerary that included two or more continents.

### Preparedness and reporting

On average, 3.73 out of the six indicators were available for the first 10 imported cases (or total number of reported importation events for countries with less than 10 total) per country (SD: 1.21, Range: 1.0-6.0). For the 21 countries with more than 10 identified importation events, 2.90 indicators were available for the subsequent 10 importation events (SD: 1.52, Range: 1.0-5.6); an average of 2.93 indicators were available for the next set of 10 importation events in the 14 countries reporting over 20 (SD: 1.72, Range: 1.0-5.7); data availability remained consistently under three average indicators for subsequent imported cases in the 11 countries reporting over 30 cases (mean = 2.80, SD: 1.42, Range: 1.0-5.2).

The median time until reporting an introduction for a country in SSA was 19 days after the first importation into the region (95% CI: 15-24 days). Time to first reported importation was significantly associated with GHSI (HR: 1.01, 95% CI: 1.00, 1.02, p=0.042), although the statistical significance of the relationship was lost after adjusting for scaled number of flight passengers per year (HR: 1.00, 95% CI: 0.99, 1.02, p=0.796). The hazard of reporting an introduction was 13.6% higher with each billion increase in the number of annual air traffic passengers (95% CI: 2.88%, 25.4%, p=0.012), after adjusting for GHSI.

Total case counts reported as of April 21st were related to GHSI. For each 10-unit increase in overall GHSI, cumulative reported case counts increased by a factor of 1.30, after controlling for the number of days since a country reported its first importation event (95% CI: 1.29-1.31), p<0.001).

### Relationship between risk of severe morbidity and death, and reporting

Assessment of within-cluster sum of squares suggested the use of five clusters for K-means analysis. Healthcare capacity factors (HCWs and beds per capita), along with the proportion of populations aged 60 and above were found to be more significantly represented within the clustering (Supplementary Figure). The clusters in Table 2 are sorted by the average proportion of the population aged 60 years and over, given its established relevance to COVID morbidity and mortality. In general, countries with larger proportions of their populations over 60 years (Clusters D and E) had higher healthcare capacity (Figure 5A), in terms of number of HCWs and hospital beds per capita. On average, cluster E countries have the largest proportion of their populations in the over 60 age range and high rates of premature death due to NCDs but also higher capacity for detecting and treating, than other clusters (Figure 5B). Despite having relatively low proportion of their populations over 60 years, countries in cluster C were considered at high risk since they tended to have fewer healthcare workers and hospital beds per capita, and a higher probability of premature death due to non-communicable diseases (NCDs) than the countries in the other four clusters (Table 2). Four of the eight countries not yet reporting cases as of March 27th (Burundi, Comoros, Lesotho, and Sierra Leone) fell into Cluster C. Both countries not reporting as of April 21st (Comoros and Lesotho are in Cluster C). Countries in this cluster that had at least one imported case did not have a significantly different data availability score for the first 10 imported cases (mean=3.56), on average, than countries in other clusters (all pairwise p > 0.05). Cluster C countries with any cases are reporting the highest case fatality ratios, on average, than countries in other clusters (4.6%, range of other CFRs: 1.5%-4.1%).

**Figure 5.**
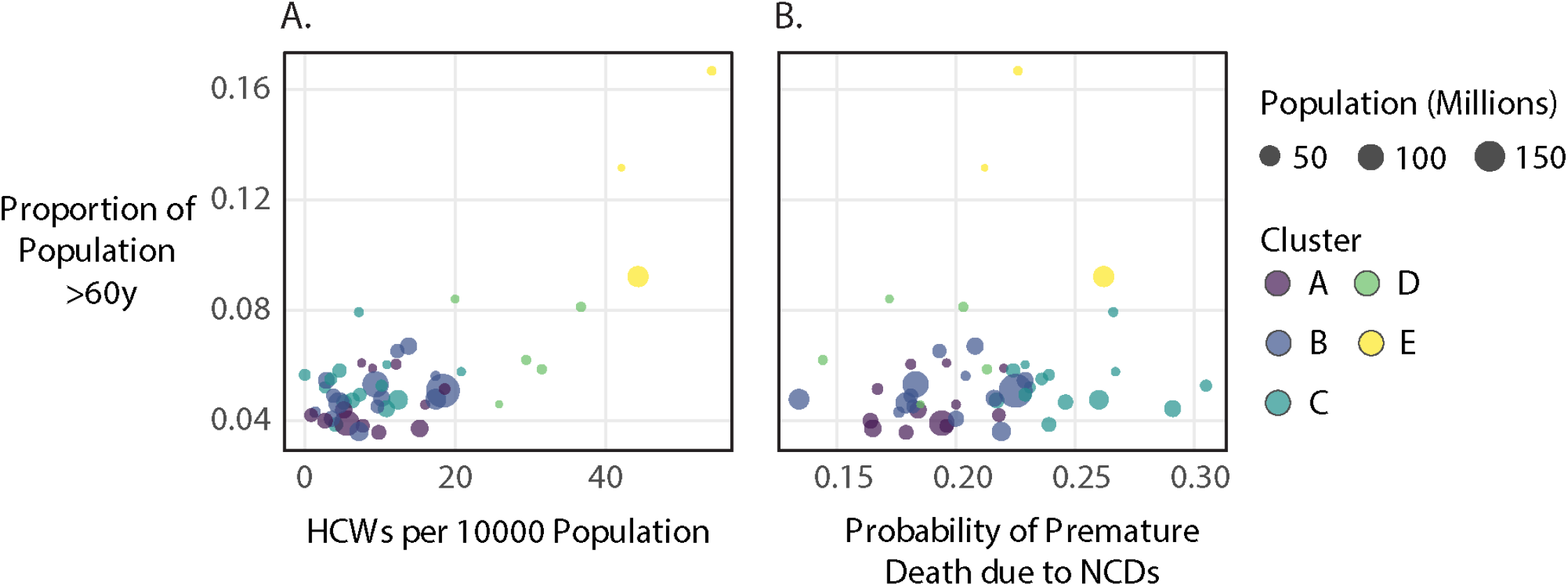
Interplay of Age, Healthcare Capacity, and Death due to Non-Communicable Disease in Sub-Saharan Africa countries. Countries with younger populations also have (A) less healthcare capacity, in terms of healthcare workers (physicians, nurses, and midwives) per 10,000 population and (B) demonstrate a wide range in terms of probability of premature death due to non-communicable comorbidities.

**Table 2.** Results of clustering analysis around risk factors for COVID-19 severe morbidity and mortality and widespread transmission of respiratory illness. Darker shades of red indicate higher risk of COVID-related morbidity and mortality relative to other clusters. Cluster characteristics (population, proportion of population aged 60+, etc. reflect average values across countries in the cluster.

| Cluster | Countries Reporting Cases | Countries Not Reporting Any Cases 30 days post-introduction | Countries Not Reporting Any Cases 54 days post-introduction | Average Population* | Prop. of Population Aged 60+ | Prob. Premature Death | HCWs | Hospital Beds | GHSI |
| --- | --- | --- | --- | --- | --- | --- | --- | --- | --- |
| A | Angola, Benin, Democratic Republic of the Congo, Djibouti, Equatorial Guinea, Guinea-Bissau, Mauritania, Mozambique, Republic of Congo (Brazzaville), Somalia, Zambia | Malawi |  | 18,274,511.83 | 0.046 | 0.19 | 8.49 | 1.04 | 24.6 |
| B | Cameroon, Ethiopia, Ghana, Kenya, Liberia, Madagascar, Niger, Nigeria, Rwanda, Senegal, Tanzania, Uganda, Zimbabwe, Gambia |  |  | 41,797,795.93 | 0.050 | 0.19 | 9.42 | 0.79 | 75.36 |
| C | Burkina Faso, Central African Republic, Chad, Côte d'Ivoire, Eritrea, Eswatini, Guinea, Mali, Sudan, Togo | Burundi, Sierra Leone | Comoros, Lesotho | 12,360,091.21 | 0.053 | 0.25 | 7.57 | 0.83 | 43.43 |
| D | Cabo Verde, Gabon, Namibia | Botswana, Sao Tome and Principe |  | 1,501,491.60 | 0.066 | 0.18 | 28.67 | 3.16 | 34.80 |
| E | Mauritius, Seychelles, South Africa |  |  | 18,403,097.67 | 0.130 | 0.23 | 46.81 | 3.27 | 79.67 |

\* Average population size for countries in each cluster

**Table 3.**
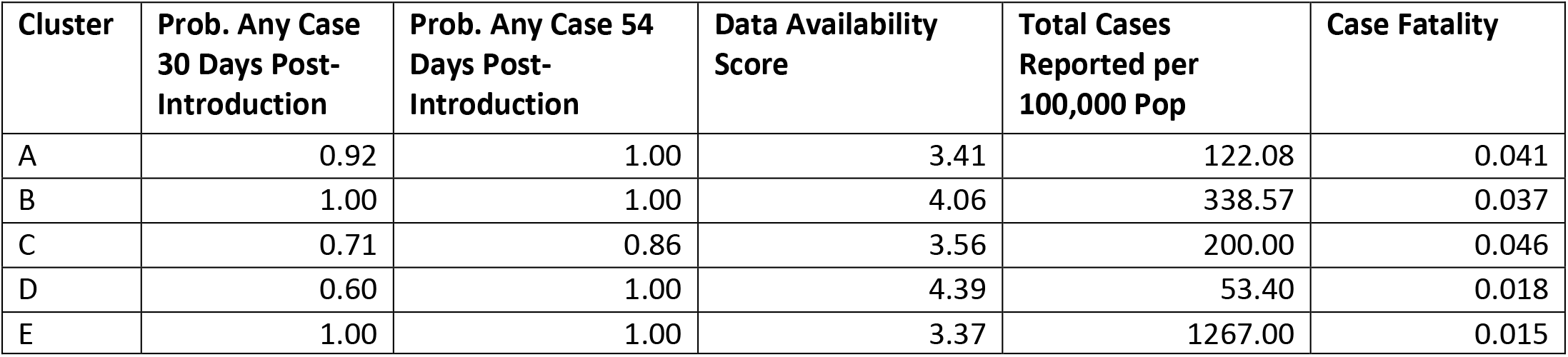
Summary of COVID-19 situation by cluster, with probability of any case reported among countries, as well average data availability scores, case counts, and case fatality ratio for countries reporting at least one case

## Discussion

We describe the seeding events that have led to over 13,200 cases reported within 54 days of the first known introduction into the sub-Saharan African region. Most documented importation events were associated with recent travel history to Europe. As of April 21st, eight countries -- Burkina Faso, Cameroon, Côte d'Ivoire, Ghana, Guinea, Niger, Nigeria, South Africa -- had reported at least 500 confirmed cases each, while two countries had reported no cases.^29^ Regional heterogeneity in reported case counts suggests that higher incidence in some places may be due in a large part to preparedness and current surveillance efforts, particularly around air travel passengers, and not solely due to more transmission. Relatedly, lower incidence in other countries may be due to reduced international air travel and the inability to surveil all air passengers and informal border crossings. Overall, importations into a country may take weeks or months to seed community spread at noticeable levels.^30^ Many importations and subsequent transmission will go unnoticed without active monitoring through quarantine and testing to identify infections among travelers^31^, and risk communication directed at population subgroups most likely interacting with international travelers to encourage vigilance around symptom onset.

In terms of assessing community spread, lack of testing capacity and decisions to test upon symptom onset for quarantined individuals^32^ could also be misconstrued for lower incidence. More worryingly, among individuals with no recent travel history, conflation of disease symptoms may lead to lack of timely health seeking and reduced access to healthcare could prove detrimental as the disease progresses and transmission increases.^33^ All of these factors could contribute to lower incidence being observed and reported. This in turn contributes to a false sense of the SSA region being less vulnerable to COVID-19 infection -- an argument that has been largely focused around age distribution alone -- when the sub-Saharan population may in fact be more vulnerable given the number of endemic, infectious and immunocompromising diseases,^16^ as well as increasing non-communicable conditions,^12^ and poor critical care infrastructure in the region.

We found that existing preparedness indicators, namely the GHSI, was associated with the number of cases reported but not with risk of earlier introduction. The sensitivity of these indicators to capturing COVID-specific differences in countries’ vulnerability to widespread transmission warrants further investigation and analysis, accounting for the different efforts being implemented across the continent to prevent disease spread.^18,21^ Our clustering analysis suggests that countries with fewer beds and healthcare workers and higher probabilities of premature death due to NCDs (thereby putting them at risk for severe COVID-19 morbidity and mortality) were less likely to report any cases during the first month post-introduction and are now reporting the highest case fatality ratios. Factors around population demographics and comorbidities uniquely relevant to COVID-19 morbidity and mortality may not be fully captured by the GHSI or other epidemic preparedness indices. The situation in SSA warrants context-specific consideration in terms of policies around timing of control efforts relative to reported case counts.

The COVID-19 situation is among the first pandemics in which high-capacity computing technology can be leveraged to quickly (and responsibly) disseminate large-scale information globally.^34,35^ Information on importation events was extracted from online reporting platforms, such as situation reports, press releases from ministries of health, and dashboards, although more often, the data were being reported via social media accounts of political leaders and governmental institutions. This reflects a growing trend intended to better connect the populations they serve with updates on the situation locally and globally.^36^

In addition to informing the general population, use of online media to share data on cases is facilitating rapid efforts at better understanding the epidemiology of the disease and its control. Notably, information from health agencies has guided mathematical modeling efforts to project case counts and assess potential impact of interventions, such as social distancing.^37-39^ The robustness of modeling efforts to inform decision-making depends on the quality of data used to develop them. Especially at the start of an outbreak, it is important to distinguish between importations and community spread among all recorded cases in order to capture the growth of the epidemic accurately.

Data needs may be overwhelming local teams, as was observed with the decreasing data availability scores with increased numbers of imported cases. This reflects competing priorities and resources, not uncommon throughout the world, and could lead to delays in providing information to decision makers. The significance of such data extends beyond information on direct consequences of COVID-19 and includes indirect consequences -- for example, if data from routine surveillance and immunization activities or care-seeking behaviors become less available to capture detrimental impacts upon routine care, as has been documented during previous large-scale outbreaks.^40–42^ Data on the burden of COVID-19, relative to the social and economic burden of control measures, are critical for informed decision-making, Support to data teams could enhance data collection so that modeling and other analyses can be done effectively and efficiently. Investing in this capacity could have long-term benefits to countries particularly those in SSA.

### Limitations

While we attempted to search all available sources, data included in our line list are likely not exhaustive. We also recognize that news sources used to supplement official sources may be less reliable in the information they provide. Information was extracted from available text material only. Videos of press conferences or radio announcements were not considered unless their content appeared in published press articles. By opting to maintain an individual-level database, we were unable to reflect some information provided by national authorities in aggregate form. Where possible, we reconciled totals across individual entries with aggregate information or adjusted for the latter, such as with the importation percentages for South Africa, Senegal, and Mauritius. Furthermore, data were not reported consistently for some indicators. For instance, we note that data being disseminated to the public are often for purposes of information sharing and education rather than to guide detailed decision making. Moreover, with increasing incidence, less information is provided publicly about cases at the individual-level, leading to lack of information on sex, age, and status (importation versus local transmission) for the majority of cases in South Africa, for instance. This precluded our investigation into whether cases with unclassified status may resemble known importations versus locally transmitted cases.

## Conclusion

The inevitable introduction of COVID-19 into sub-Saharan Africa has led to variation in the incidence and reported ratio of imported versus locally infected cases. The availability and quality of publicly released information also varies significantly. Countries with systemic, demographic, and pre-existing health vulnerabilities to severe COVID-related morbidity and mortality are less likely to report any cases and some are reporting with limited public availability of information. Lack of information on imported cases should signal the potential for undetected transmission with consequential direct and indirect implications for the population and healthcare systems in these countries.

## Data Availability

The data are freely available and are included as a supplement.

## Acknowledgments

We acknowledge Kurt Frey for help in collecting data and Assaf Oron for helpful comments on the manuscript. We also acknowledge the tireless efforts of country teams to collect and disseminate information as a way to inform local and global stakeholders.

## Author contributions

LAS, PS, BH, ALO, NN, and DM collected data. LAS conducted the analysis. LAS, PS, and BMA wrote the first draft of the manuscript. All authors contributed intellectually and made contributions to the manuscript text.

## Ethics approval and consent to participate

This study uses publicly available data thus ethical approval does not apply.

## Availability of data and materials

Data is available with the manuscript.

